# IL-6 and IL-10 as predictors of disease severity in COVID-19 patients: results from meta-analysis and regression

**DOI:** 10.1101/2020.08.15.20175844

**Authors:** Sujan K Dhar, K Vishnupriyan, Sharat Damodar, Shashi Gujar, Manjula Das

## Abstract

**Aims:** SARS-CoV-2, an infectious agent behind the ongoing COVID-19 pandemic, induces high levels of cytokines such as IL-1, IL-2, IL-4, IL-6, IL-10, TNF-α, IFN-γ etc in infected individuals which contribute towards the underlying disease patho-physiology. Nonetheless, exact association and contribution of every cytokine towards COVID-19 pathology remains poorly understood. Delineation of the role of the cytokines during COVID-19 holds the key of efficient patient management in clinics. This study performed a comprehensive meta-analysis to establish association between induced cytokines and COVID-19 disease severity to help in prognosis and clinical care.

**Main methods:** Scientific literature was searched to identify 13 cytokines (IL-1β, IL-2, IL-2R, IL-4, IL-5, IL-6, IL-7, IL-8, IL-10, IL-12, IL-17, TNF-α and IFN-γ) from 18 clinical studies. Standardized mean difference (SMD) for selected 6 cytokines IL-2, IL-4, IL-6, IL-10, TNF-α and IFN-γ between severe and non-severe COVID-19 patient groups were summarized using random effects model. A classifier was built using logistic regression model with cytokines having significant SMD as covariates.

**Key findings:** Out of 13 cytokines, IL-6 and IL-10 showed statistically significant SMD across the studies synthesized. Classifier with mean values of both IL-6 and IL-10 as covariates performed well with accuracy of ~ 92% that was significantly higher than accuracy reported in literature with IL-6 and IL-10 as individual covariates.

**Significance:** Simple panel proposed by us with only two cytokine markers can be used as predictors for fast diagnosis of patients with higher risk of COVID-19 disease deterioration and thus can be managed well for a favourable prognosis.

## Introduction

COVID-19 pandemic caused by SARS-CoV-2 has emerged as a major threat to mankind affecting 18+ million people resulting in over 700,000 deaths (1) worldwide. Exuberant inflammation manifested as elevated levels of cytokines, commonly referred as “cytokine storm” (CS) often leads to critical conditions like ARDS (acute respiratory distress syndrome) and death due to multi-organ failure(2).

Innate immune response is the first step of defence mechanism against viral infection. Pattern recognition receptors in host dendritic cells recognize viral genomic DNA or RNA to initiate production of cytokines and chemokines(3) which in turn recruit immune cells like macrophages, neutrophils and T-cells to the site of infection based on their source and target cells(4). Pro-inflammatory cytokines including type I/II interferons, IL-1, IL-6 and TNF-α play major role in initial response, whereas anti-inflammatory molecules like IL-10 are produced during sustained infection to keep a check on inflammation and maintain immune homeostasis(5). Heightened CS induced acute lung damage leading to fatality(6) is a signature of coronavirus family reported earlier for MERS-CoV and SARS-CoV infections(7,8).

In COVID-19, elevated levels of both pro-inflammatory and anti-inflammatory cytokines are reported in multiple clinical studies (9-11). A recently published extensive meta-analysis summarized elevated levels of IL-2, IL-2R, IL-4, IL-6, IL-8, IL-10, TNF-α and IFN-γ in severe group of patients, whereas no significant increase was found in the levels of IL-1β and IL-17(12). A two-arm meta-analysis study synthesized from individual patient data reported statistically significant odds ratio (p < 0.05) to sever disease for only two cytokines – IL-6 and Il-10(13). Some meta-analysis efforts reported difference in IL-6 level between severe and non-severe COVID-19 patients in terms of Standardized Mean Difference (SMD) (14), mean difference (15) or ratio of means (16) values that can potentially be used as cut-off to discriminate between severe and non-severe patients. All these meta-analysis summary results were associated with high level of heterogeneity (I^2^ ~ 98 – 100%). Another synthesis over three clinical studies determined the elevated ratio IL-6/IFN-γ in severe patients (17) with substantial heterogeneity (I^2^ = 79%).

Clinical studies with COVID-19 patient cohorts explored the role of IL-6 alone (18) or along with other cytokines including IL-10, IL-2, IL-4, TNF-α and IFN-γ as prognosticator for severe disease(19). Meta-analysis studies described before(12,14-16,20,21) concluded elevated levels of cytokines in severe COVID-19 patients but did not attempt to establish their prognostic significance, except the study by Elshazli et al who performed decision tree and ROC curve analysis to assess prognostic potential of multiple laboratory parameters including IL-6(13). However, despite numerous clinical studies and meta-analyses any reliable prognostic method that can predict the progression of a patient to severe form of disease based on on-admission cytokine levels remains elusive. In this study, we attempt to arrive at a prognostic method through meta-analysis of the levels of commonly used 13 cytokine markers between severe and non-severe patient groups by building a classifier using a logistic regression model.

## Methods

### Literature Search

We performed the meta-analysis following PRISMA guidelines(22). Literature search was performed in Pubmed, Google Scholar and in preprint archives such as medRxiv, bioRxiv and SSRN library for articles in English published in year 2020 till 31 May 2020. Search terms included COVID-19-specific terms in title of article (“2019-nCov” OR “nCoV-2019” OR “novel coronavirus” OR “SARS-CoV-2” OR “COVID-19” OR COVID19 OR “novel corona virus”) along with terms such as “cytokine level” and combinations of common cytokine names and gene symbols. Search strategy was reviewed by all authors and it was decided to retain non-peer reviewed articles in selection in view of the present emerging situation. Identified articles were screened and shortlisted to clinical studies with inclusion criteria as only clinical studies with laboratory data for at least two cytokines for severe and non-severe COVID-19 patient groups. Exclusion criteria for shortlisting included review articles, opinions and commentaries, studies that include other pathological conditions or complications associated with COVID-19 and studies without mean (SD) or median (IQR) data of cytokines for each group.

### Data Extraction

Selected articles were reviewed independently by two authors (VK and SKD). Data was extracted from articles using standardized forms and was cross-validated. Cytokine levels reported as median (IQR) was converted to mean (SD) using standard methods(23). One study reported non-severe patient data across two groups(24) which were combined using recommendations in Cochrane Handbook(25) section 6.5.2.10. Another study represented data in subgroups of IgG levels and Neutrophil-to-Lymphocyte ratio (NLR) (26), for which we retained data from only the IgG-low subgroups as that would be closest to early stage of infections, and combined subgroups of high and low NLR values.

### Meta-Analysis and Meta-Regression

To assess the effect of each marker, we used the standardized mean difference (SMD) of measured cytokine level (in pg/mL) between severe and non-severe groups including Hedges’ correction for positive bias. All calculations were carried out using the metafor library(27) on R statistical software platform. Meta-regression of SMD of a marker was carried out using mixed-effects model with differences in age and sex (measured as percentage of male patients). Publication bias in the studies was analyzed using Funnel Plot and Egger’s regression test. Sensitivity of meta-analysis results was assessed by repeating the analysis with leaving out one study at a time and observing significant changes in heterogeneity and summary effect.

### Classifier Development

We built a classifier using logistic regression model for categorization of patient groups in to severe and non-severe categories based on mean values of cytokines in the groups. Classifier performance using individual cytokine values or combination of multiple cytokine values are assessed from the Receiver Operating Characteristics (ROC) curve analysis and the area under the curve (AUC).

## Results

### Study Details

The search yielded a total of 99 “hits” which were screened to select 18 articles containing cytokine levels of severe and non-severe patients reported from clinical studies (Fig. 1). All 18 selected studies were conducted in China and included 1,242 non-severe and 915 severe COVID-19 patients (Table 1). Out of the 18 studies, 7 were non-peer reviewed studies(24,26,28-32) published online in preprint archives.

**Figure 1:**
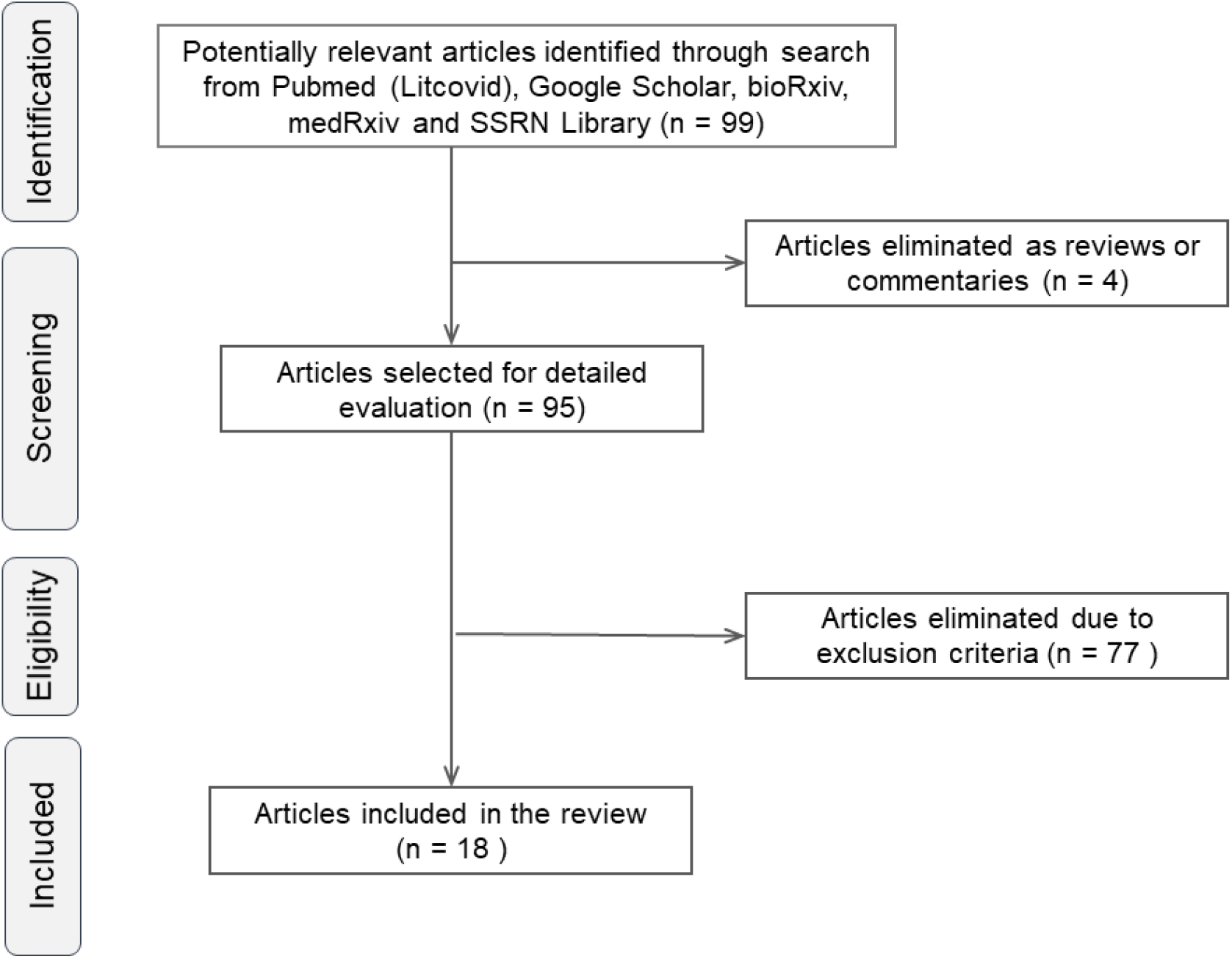
Workflow for selection of studies.

**Table 1:** Details of selected studies.

| Author<br>(year) | Non-severe patients |  |  | Severe Patients |  |  | Clinical<br>Definitions of<br>Severe Patients | Cytokines<br>Measured |
| --- | --- | --- | --- | --- | --- | --- | --- | --- |
|  | n | Age | Male<br>% | n | Age | Male<br>% |  |  |
| Chen,<br>2020(33) | 10 | 50.3<br>(10.1) | 70.0 | 11 | 61.2<br>(7.3) | 90.9 | NHC guideline,<br>pulmonary<br>lesion | IL-6, IL-10,<br>TNF-α |
| Feng,<br>2020(28) | 94 | 62.9<br>(13.7) | 61.7 | 20 | 69.2<br>(11.1) | 65.0 | NHC guideline | IL-2, IL-4,<br>IL-6, IL-10,<br>TNF-α, INF-γ |
| He,<br>2020(34) | 12 | 42.3<br>(16.8) | 31.1 | 16 | 62.3<br>(16.7) | 53.6 | NHC guideline | IL-2, IL-4,<br>IL-6, IL-10,<br>TNF-α, INF-γ |
| Lv,<br>2020(35) | 115 | 54.3<br>(41.5) | 50.4 | 155 | 58.7<br>(47.4) | 49.7 | Not specified | IL-2, IL-4,<br>IL-6, IL-10,<br>TNF-α, INF-γ |
| Nie,<br>2020(24) | 72 | 40.3<br>(19.3) | 29.1 | 25 | 57.3<br>(15) | 52.0 | NHC guideline,<br>pulmonary<br>lesion | IL-2, IL-4,<br>IL-6, IL-10,<br>TNF-α, INF-γ |
| Qin,<br>2020(36) | 166 | 52.1<br>(15.4) | 48.2 | 286 | 57<br>(13.3) | 54.2 | NHC guideline | IL-6, IL-10,<br>TNF-α |
| Shi,<br>2020(29) | 22 | 49.7<br>(12) | 59.7 | 21 | 59<br>(17.3) | 72.4 | NHC guideline,<br>pulmonary<br>lesion | IL-2, IL-4,<br>IL-6, IL-10,<br>TNF-α, INF-γ |
| Song,<br>2020(30) | 31 | 48<br>(16.5) | 51.6 | 42 | 55.9<br>(12.2) | 71.4 | NHC guideline,<br>pulmonary<br>lesion | IL-2, IL-4,<br>IL-6, IL-10,<br>TNF-α, INF-γ |
| Wan,<br>2020(37) | 102 | 43.1<br>(13.1) | 53.9 | 21 | 61.3<br>(15.6) | 52.4 | NHC guideline | IL-4, IL-6,<br>IL-10, TNF-α,<br>INF-γ |
| Wang,<br>2020(38) | 55 | 40<br>(14.1) | 45.0 | 14 | 69.8<br>(11.4) | 50.0 | NHC guideline<br>with SpO<sub>2</sub> <<br>90% | IL-2, IL-4,<br>IL-6, IL-10,<br>TNF-α |
| Wei, Su,<br>2020(39) | 131 | 60.1<br>(12.4) | 45.0 | 98 | 69.7<br>(11.6) | 60.0 | NHC guideline,<br>pulmonary<br>lesion | IL-2, IL-4,<br>IL-6, IL-10,<br>TNF-α, INF-γ |
| Xu,<br>2020(40) | 80 | 55.7<br>(17) | 37.5 | 45 | 57.7<br>(15.7) | 68.9 | NHC guideline | IL-6, IL-10 |
| Zhang, Zhou,<br>2020(26) | 44 | 53.5<br>(20.1) | 47.7 | 21 | 58.3<br>(13.9) | 71.4 | Not specified | IL-2, IL-4,<br>IL-6, IL-10,<br>TNF-α, INF-γ |
| Zhang,<br>Wang,<br>2020(31) | 29 | 44.3<br>(15.8) | 58.6 | 14 | 61.7<br>(9.2) | 35.7 | NHC guideline | IL-2, IL-4,<br>IL-6, IL-10,<br>TNF-α, INF-γ |
|  | n | Age | Male<br>% | n | Age | Male<br>% |  |  |
| Zhang, Yu,<br>2020(41) | 93 | 38.2<br>(12.2) | 34.4 | 18 | 63.3<br>(24.9) | 77.8 | WHO guideline | IL-2, IL-4,<br>IL-6, IL-10,<br>TNF-α, INF-γ |
| Zhao,<br>2020(32) | 53 | 44.8<br>(15.9) | 43.4 | 18 | 63.9<br>(17.2) | 38.9 | NHC guideline | IL-2, IL-4,<br>IL-6, IL-10,<br>TNF-α, INF-γ |
| Zheng,<br>2020(42) | 22 | 46.9<br>(14.7) | 40.9 | 74 | 56.8<br>(13.7) | 66.2 | NHC guideline | IL-2, IL-4,<br>IL-6, IL-10,<br>TNF-α, INF-γ |
| Zhu,<br>2020(43) | 111 | 50<br>(15.5) | 65.8 | 16 | 57.5<br>(11.7) | 56.3 | NHC guideline,<br>pulmonary<br>lesion | IL-2, IL-4,<br>IL-6, IL-10,<br>TNF-α, INF-γ |

NHC guideline: (a) Respiratory distress, $RR \geq 30$ /min, (b) Oxygen saturation level ( $SpO_2$ ) $\leq 93\%$ at rest and (c) Arterial blood oxygen partial pressure ( $PaO_2$ ) /oxygen concentration ( $FiO_2$ ) $\leq 300$ mm Hg.
Pulmonary lesion: $>50\%$ lesion progression within 24-48h in pulmonary imaging
Age in mean (SD)

In most studies the severe patient group was designated based on National Health Commission of China guidelines(44) that specifies (a) Respiratory distress, RR≥30/min, (b) Oxygen saturation level (SpO_2_) ≤ 93% at rest and (c) Arterial blood oxygen partial pressure (PaO_2_) /oxygen concentration (FiO_2_) ≤300 mm Hg. For some studies, pulmonary lesion progression > 50% within 24-48 hours on imaging was used as additional criteria. One study used WHO guideline(45) that categorizes severe patients as adolescent or adult with clinical signs of pneumonia (fever, cough, dyspnoea, fast breathing) plus one of the following: respiratory rate > 30 breaths/min; severe respiratory distress; or SpO_2_ < 90% on room air.

Weighted mean of age of patients in severe group (59.7y, range 55.9 – 69.8) was higher than the non-severe group (50.2y, range 38.2 – 62.9). Severe group also had larger percentage of male patients (57.8%, range 35.7 – 90.9) than non-severe group (48.4%, range 29.1 – 70.0).

Measurement of total 13 cytokines (IL-1β, IL-2, IL-2R, IL-4, IL-5, IL-6, IL-7, IL-8, IL-10, IL-12, IL-17, TNF-α and IFN-γ) were reported in the selected 18 studies. However, 13 studies were considered for further analysis since they reported analysis of five or more studies in at least 1000 patients (Supplementary Table S1). These studies reported levels of only 6 cytokines (IL-2, IL-4, IL-6, IL-10, TNF-α and IFN-γ).

### Meta-analysis of Cytokine Levels

Meta-analysis for SMD value of each marker using a random-effects model showed moderate and statistically significant elevation in severe patients for two cytokines viz. IL-6 (SMD 0.53, 95% CI: 0.26 – 0.80, p < 0.001) (Fig. 2a) and IL-10 (SMD 0.64, 95% CI: 0.38 – 0.91, p < 0.0001) (Fig. 2b). Level of IFN-γ, the type II interferon, showed a weak elevation in severe group (SMD 0.11, 95% CI: −0,01 – 0.23, p = 0.078) with moderate significance. Summary effect size of other markers IL-2 (p = 0.281), IL-4 (p = 0.305) and TNF-α (p = 0.258) were not significant (Table 2).

**Figure 2:**
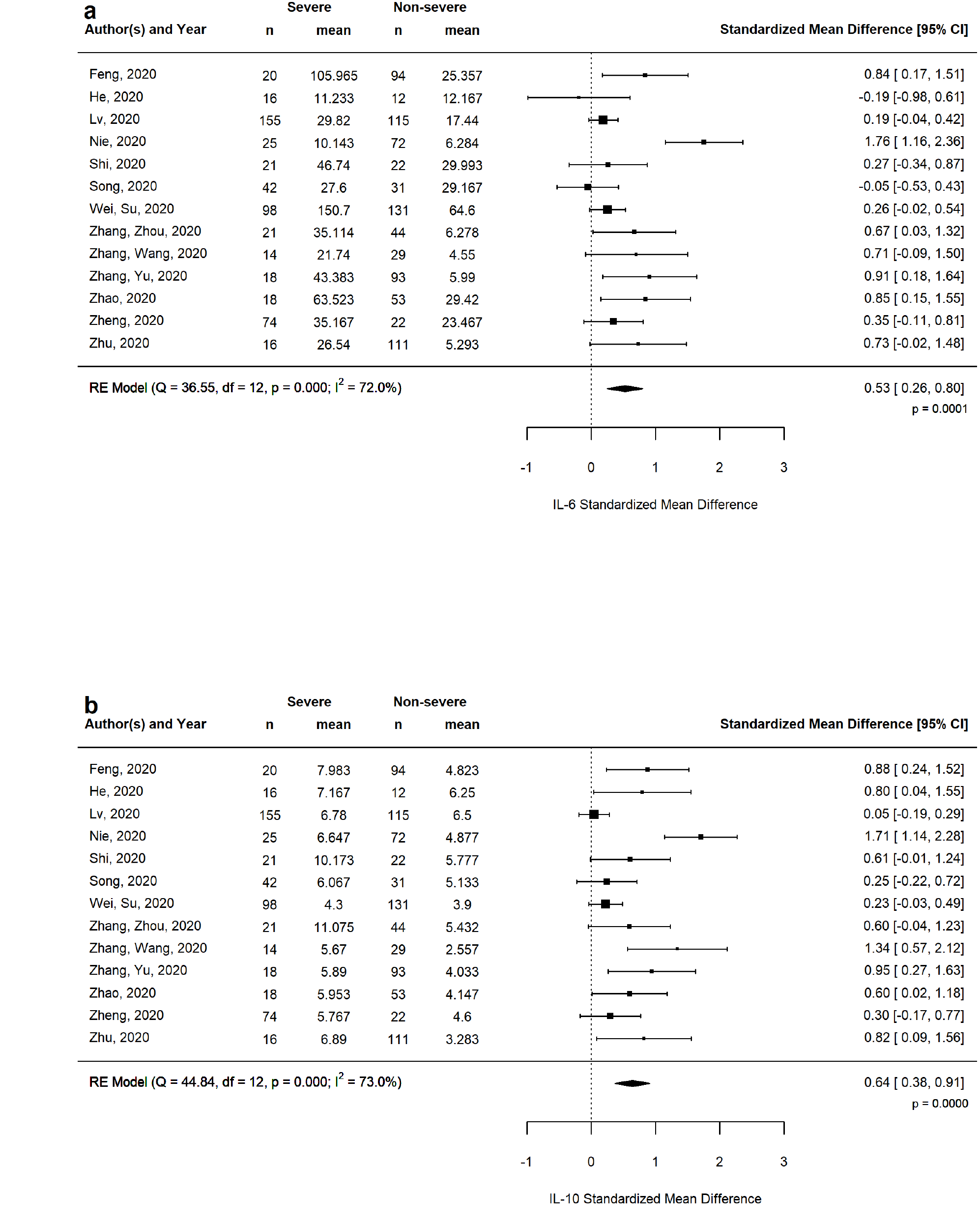
Standardized mean difference of (a) IL-6 and (b) IL-10 between severe and non-severe COVID-19 patients.

**Table 2:** Meta-analysis result summary of six cytokines.

| Cytokine | Random Effect Model SMD<br>(95% CI) | Summary Effect<br>p.value | Heterogeneity (I<sup>2</sup>) |
| --- | --- | --- | --- |
| IL-2 | 0.12 (-0.10, 0.35) | 0.281 | 66.9% |
| IL-4 | 0.15 (-0.13, 0.42) | 0.305 | 79.2% |
| **IL-6** | **0.53 (0.26, 0.80)** | **0.0001** | **72.0%** |
| **IL-10** | **0.65 (0.38, 0.91)** | **<0.0001** | **73.0%** |
| TNF-α | 0.10 (-0.08, 0.28) | 0.258 | 50.8% |
| IFN-γ | 0.11 (-0.01, 0.23) | 0.078 | 0% |

### Meta-Regression with Age and Gender Difference

Summary effects of both IL-6 and IL-10 showed substantial heterogeneity (I^2^ ~ 72-73%) source of which was explored by carrying out meta-regression of IL-6 and IL-10 using (i) SMD of patient age and (ii) difference in male percentage between severe and non-severe groups as moderators in a mixed-effects model. Results revealed IL-10 levels having a dependence on the age difference (coefficient 0.63, p = 0.012), whereas the dependence of IL-6 on age was not significant (p > 0.05, Fig. 3). No dependence of IL-6 and IL-10 levels were found on gender expressed as difference of male percentage between the two groups (Supplementary Fig. S1). Above results ascribe some of the observed heterogeneity in IL-6 and IL-10 SMD to difference in average age between severe and non-severe group.

**Figure 3:**
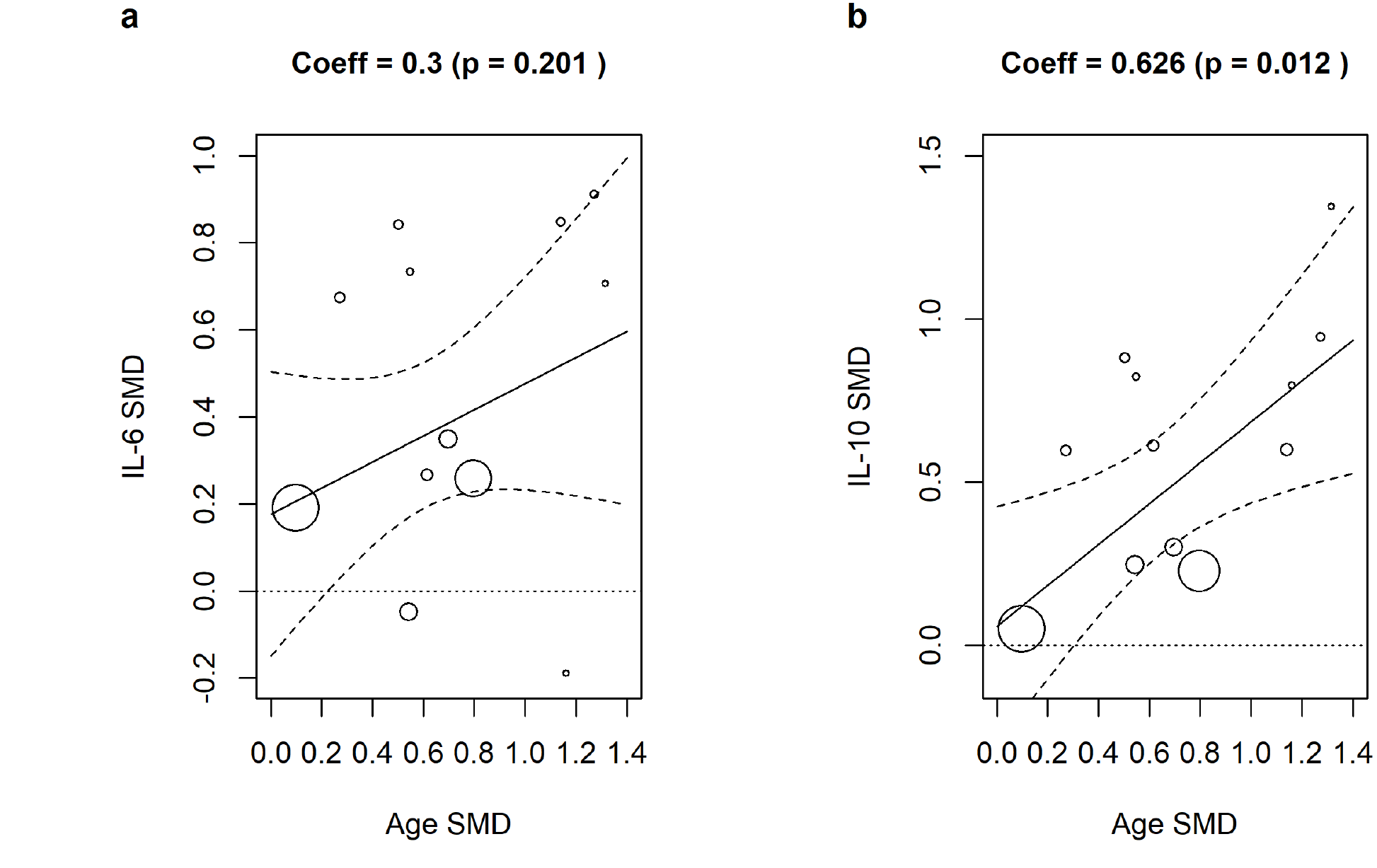
Meta-regression bubble plots of (a) IL-6 and (b) IL-10 SMD with difference in age between severe and non-severe groups.

### Classifier Performance

Our results indicate that out of the analysed cytokines, levels of only the pro-inflammatory IL-6 and the immuno-suppressive IL-10 are significantly elevated in the severe group of patients, an observation reported in other studies as well(19,46). This prompted us to assess the prognostic potential of these markers by developing a logistic regression model using IL-6 and IL-10 mean values as covariates to classify the patient groups as severe and non-severe across all 18 studies. Model using both IL-6 and IL-10 as covariates showed an overall accuracy of 91.7% and area under the corresponding ROC curve as 0.957 (95% CI: 0.898 – 1), which is higher than the corresponding values using IL-6 (accuracy 77.8%, AUC 0.821) and IL-10 (accuracy 80.6%, AUC 0.878) individually as covariates (Table 3, Fig. 4). Model using both IL-6 and IL-10 showed nearly 100% specificity and 83.3% sensitivity for classification in severe and non-severe categories.

**Table 3:**
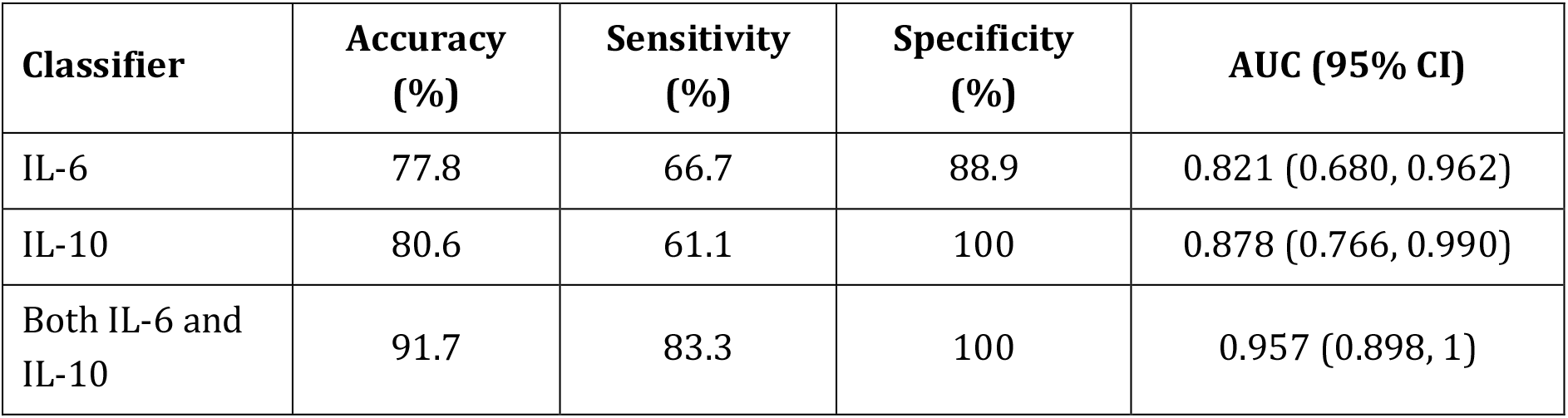
Classifier performance summary using only IL-6, only IL-10 and both IL-6 and IL-10 as input variables.

| Classifier | Accuracy (%) | Sensitivity (%) | Specificity (%) | AUC (95% CI) |
| --- | --- | --- | --- | --- |
| IL-6 | 77.8 | 66.7 | 88.9 | 0.821 (0.680, 0.962) |
| IL-10 | 80.6 | 61.1 | 100 | 0.878 (0.766, 0.990) |
| Both IL-6 and IL-10 | 91.7 | 83.3 | 100 | 0.957 (0.898, 1) |

**Figure 4:**
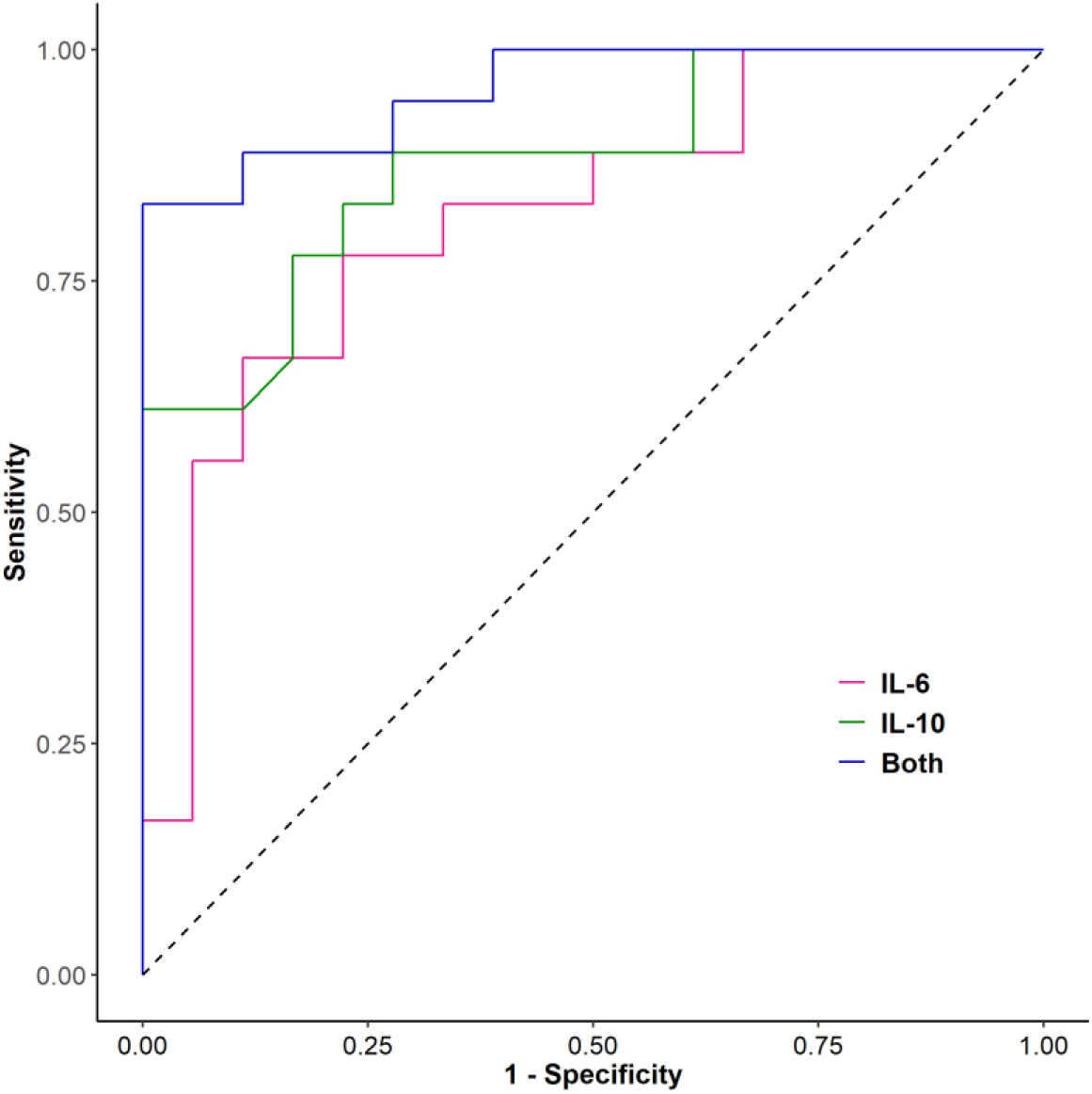
ROC curve for classifier performance using only IL-6, only-10 and both IL-6 and IL-10 as input parameters.

### Publication Bias and Sensitivity

Publication bias towards higher SMD of IL-10 levels reported in smaller-size studies manifested as asymmetry in Funnel plot and Egger’s regression test (p < 0.001), whereas the corresponding bias was not significant (p = 0.151) for IL-6 (Supplementary Fig. S2). Sensitivity analysis of the studies showed a large reduction of heterogeneity in IL-6 (I^2^ value from 72% to 25%) and a moderate reduction for IL-10 (73% to 55%) by leaving out one particular study from the analysis (Supplementary Fig. S3).

### Limitations

Primary limitation of this meta-analysis is that the studies are restricted only to Chinese ethnicity, which was not by design but due to the fact that most initial studies were reported from this geographic region, the site of first spread. Also 7 of the 18 studies were non-peer reviewed publications selected as per search strategy. There was some heterogeneity in clinical criteria for severe disease as shown in Table 1. Also recruitment of patients in the studies were at different stages of disease progression, which might have caused bias in the on-admission measurement of laboratory parameters in cytokines.

## Discussion

With the sustained spread of COVID-19 in most societies, a suitable prognostic test that can predict progression of a patient to severe state of disease with reasonable accuracy is need of the hour for efficient management and care. Cytokine storm caused by a dysregulation of immune system through increase in immune cell activation versus suppression of anti-inflammatory response is likely to hold the key in discovery of such prognostic factors.

Elevated levels of IL-6, a pro-inflammatory molecule, is known to down-modulate NK cell activity and are also found to be associated with reduction in granzyme and perforin levels causing impairment of lytic activities(47). In COVID-19 patients exacerbation symptoms such as increased body temperature, elevation in inflammation markers like CRP and serum ferritin and progressed chest computed tomography images were associated with increased IL-6 levels which showed a downturn during recovery(48). This association of IL-6 with pulmonary conditions were reported earlier in patients with pneumonia(49) or severe pneumonitis caused by radiation therapy(50).

IL-10, on the other hand, is an anti-inflammatory cytokine that was found elevated in severe COVID-19 patients(2,33,36). Levels of IL-6, IL-10 and TNF-α were also found to be indicators of T-cell exhaustion in COVID-19 patients(51). IL-10 is a multifunctional cytokine whose primary function is to limit the inflammatory response. However, IL-10 is also known to introduce anergy or non-responsiveness of T-cells in anti-tumour cell response(52) as well as in viral infection(53). Blockade of IL-10 using antibody against IL-10 or its receptor or genetic removal of IL-10 resulted in elimination of infection by virus(53-55) or bacterial pathogen(56).

Though the levels of cytokines are increased in severe COVID-19 patients, it’s implication in a therapeutic perspective remains unclear. Corticosteroids that can potentially suppress cytokines by inhibiting NF-kB transcription factor have been used on COVID-19 patients. However, a meta-analysis across studies involving infection of SARS-CoV, SARS-CoV-2 and MERS-CoV showed increased mortality risk ratio (RR 2.11, 95% CI: 1.13 – 3.94) for patients treated with corticosteroids(57). In an unique study, anti-TNF-α antibody drug, infliximab, was administered to a small group (n = 7) of severe COVID-19 patients without any pre-existing auto-immune condition(58). Though the patients showed decrease in IL-6 level and increase in lymphocyte count post-administration, the cohort size was not adequate to make any conclusion on the end-point benefits. Few other studies used etanercept(59), adalimumab(60) and infliximab(61) therapy successfully for COVID-19 patients, but with pre-existing auto-immune conditions. On the other hand, there have been several studies targeting IL-6 levels using siltuximab, an IL-6 inhibitor or tocilizumab, an IL-6 receptor inhibitor in severe COVID-19 patients. A recent review (62) on few such studies did not reveal any conclusive benefits due to lack of appropriate control groups in design or inadequate statistical significance. Further, the timing for anti-IL-6 agent administration remains critical since an early administration may negatively affect the innate response, while the late administration may not yield desired benefits. Some of the ongoing clinical results using anti-IL-6 therapy will shed more light in near future.

Therapeutic strategy of targeting IL-10 has been a nascent topic for infective diseases. Mice models treated with the immunomodulator compound Ammonium trichloro(dioxyethylene-O-O’)tellurate (AS101) that inhibits IL-10 transcription showed an improvement in survival from sepsis-related death(63). Similarly, lymphocytic choriomeningitis virus (LCMV) infected mice when treated with anti-IL-10 antibody exhibited enhanced T-cell responses resulting in increased IFN-γ production and reduction in viral load(53). Blockade of IL-10 signalling using an antibody that targets IL-10 receptor also reduced viral titre and persistence in LCMV-infected mice(54,55). Similar in vivo experiments with SARS-CoV-2 infected animal models will be needed to establish IL-10 as a therapeutic target for severe and critical COVID-19 disease.

Despite the unclarity on therapeutic potential of IL-6 and IL-10, their observed levels of elevation in severe COVID-19 patients have prompted clinical researchers to explore use of them as prognosticators. In an earlier study with pneumonia-affected children, IL-6/IL-10 ratio on admission was found to be an indicator of severe disease with sensitivity and specificity of 76.5% and 83.3% respectively (49). In COVID-19 context, ROC curve analysis on an Italian cohort of 77 patients showed IL-6 as a prognostic factor for combined end-point of progression to severe disease and/or in-hospital mortality with AUC = 0.8(18). However, performance of prognosis for progression to severe disease was poorer with AUC = 0.75. In the recently reported meta-analysis over a large number of studies, classifier performance was found to be higher for neutrophil count (AUC = 0.831), lymphocyte (AUC = 0.867) and D-dimer (AUC = 0.876) than using IL-6 as covariate(AUC = 0.632)(13). Another research with 102 COVID-19 patients used logistic regression to evaluate potential of multiple cytokines to diagnose severe and critical patients. Results revealed best prognostic performance for IL-6 (AUC = 0.83), followed by IL-10 (AUC = 0.73) as individual covariates whereas the model with all cytokines showed marginally better performance (AUC 0.86, sensitivity 80.0%, specificity 75.9%).

Other than these cytokines, D-Dimer, CRP level and NLR are explored as diagnostic or prognostic markers in COVID-19. Though a couple of studies focused only on their diagnostic potential for COVID-19 positivity(64,65), ROC analysis in a study with 84 patients hospitalized with COVID-19 pneumonia(66) showed modest performance of NLR and CRP to predict a 7-day endpoint as AUC values of 0.69 (CRP), 0.76 (NLR) and 0.080 (both). Another study with 93 confirmed patients established a slightly higher performance with AUC values as 0.84(NLR) and 0.71(CRP) to distinguish severe and non-severe patients. Pooled ROC curve analysis in a couple of meta-analysis studies showed modest classifier performance for CRP and D-Dimer with AUC values as 0.84 and 0.69(67) and 0.88 and 0.77(68) respectively to discriminate severe and non-sever patients. Since in all these cases the AUC values are below 0.9, predictive values for NLR, CRP and D-Dimer to diagnose severe patients may remain restricted. A recent proteomics study(69) predicted excellent classification performance (AUC = 0.96) for severe and non-severe COVID-19 patients using 22 serum proteins and 7 metabolites as covariates. However, a proteomics infrastructure is not so common in a clinic, limiting clinical utility of this test.

In this context, results of the meta-analysis and classification obtained in this study assumes importance in multiple aspects. Firstly, synthesis of a large number of studies (more than many of the cited meta-analyses) demonstrates a significant difference in values for only two cytokines – IL-6 and IL-10 with counter-acting mechanisms, reaffirming the dysregulation of immune response in progression to a severe state. Secondly, even at the absence of individual patient data, ROC curve analysis with mean values of IL-6 and IL-10 levels manifests a high level of classifier performance (AUC > 0.9) in distinguishing severe from non-severe group patients, which is higher than classifier performance from individual studies(18,19). Our results are likely to be more robust since variations from multiple patient cohorts are included in this synthesis. And lastly, it opens the possibility of further research to establish a validated diagnostic test using these two markers for management of COVID-19 patients. Since we are working with mean values of markers in severe or non-severe patient groups and not individual patient-level values, we have refrained from using IL-6/IL-10 ratio as the classifier as was done in the pneumonia study(49). However, availability of patient data from multiple cohorts of severe and non-severe patients can firmly establish and validate the test with the ratio or any other transform as the basis of classification.

## Conclusion

Our findings, summarized over multiple studies, indicate a possible dysregulation in immune response against COVID-19, characterized by two counter-acting cytokines IL-6 and IL-10, shifts the balance between non-severe to severe category of patients, and hence measurement of both markers is necessary to demarcate the boundary. Measurement of serum levels IL-6 and IL-10 are inexpensive and can be performed on admission to clinics or care centres with minimum facilities. Such measurement will be key to identify patients with a greater likelihood of progression to severe disease and thereby to adopt necessary precautionary measures.

Further trials with multiple cohorts of COVID-19 patients or assimilation of patient-level data from existing cohorts will be needed to establish and validate the test. Other than the diagnostic potential, option of possible therapeutic strategy targeting either IL-6 or IL-10 or both is likely to emerge through analysis of such data.

## Data Availability

All data referred in this manuscript are available in the manuscript itself or in supplementary data.

## Author Contributions

SKD: Conceptualization, Data curation, Formal analysis, Writing – original draft, VK: Data curation, Writing – original draft, SD: Validation, SG: Writing – editing & review, MD: Project administration, Writing – editing & review

## Acknowledgement

SKD acknowledges Prof. Joy Kuri, Chair, Department of Electronic Science and Engineering, Indian Institute of Science, Bangalore for providing the computational resources.

## Supplementary Material

Enclosed

## References

1. Coronavirus Disease (COVID-19) Situation Report – 199. World Health Organization. 2020.

2. Huang C, Wang Y, Li X, Ren L, Zhao J, Hu Y, et al. Clinical features of patients infected with 2019 novel coronavirus in Wuhan, China. Lancet. 2020 Feb;395(10223):497–506.

3. Kawai T, Akira S. Innate immune recognition of viral infection. Nat Immunol. 2006;7(2):131–7.

4. Turner MD, Nedjai B, Hurst T, Pennington DJ. Cytokines and chemokines: At the crossroads of cell signalling and inflammatory disease. Biochim Biophys Acta – Mol Cell Res. 2014;1843(11):2563–82.

5. Rojas JM, Avia M, Martín V, Sevilla N. IL-10: A multifunctional cytokine in viral infections. J Immunol Res. 2017;2017.

6. Tisoncik JR, Korth MJ, Simmons CP, Farrar J, Martin TR, Katze MG. Into the Eye of the Cytokine Storm. Microbiol Mol Biol Rev. 2012;76(1):16–32.

7. Lau SKP, Lau CCY, Chan KH, Li CPY, Chen H, Jin DY, et al. Delayed induction of proinflammatory cytokines and suppression of innate antiviral response by the novel Middle East respiratory syndrome coronavirus: Implications for pathogenesis and treatment. J Gen Virol. 2013 Dec;94(PART 12):2679-90.

8. Channappanavar R, Perlman S. Pathogenic human coronavirus infections: causes and consequences of cytokine storm and immunopathology. Semin Immunopathol. 2017;39(5):529–39.

9. Ragab D, Salah Eldin H, Taeimah M, Khattab R, Salem R. The COVID-19 Cytokine Storm; What We Know So Far. Front Immunol. 2020;11(June):1-4.

10. Costela-Ruiz VJ, Illescas-Montes R, Puerta-Puerta JM, Ruiz C, Melguizo-Rodriguez L. SARS-CoV-2 infection: The role of cytokines in COVID-19 disease. Cytokine Growth Factor Rev. 2020;(May):0-1.

11. Tan M, Liu Y, Zhou R, Deng X, Li F, Liang K, et al. Immunopathological characteristics of coronavirus disease 2019 cases in Guangzhou, China. Immunology. 2020;160(3):261–8.

12. Akbari H, Tabrizi R, Lankarani KB, Aria H, Vakili S, Asadian F, et al. The role of cytokine profile and lymphocyte subsets in the severity of coronavirus disease 2019 (COVID-19): A systematic review and meta-analysis. Life Sci. 2020;258(July):118167.

13. Elshazli R, Toraih EA, Elgaml A, El-Mowafy M, El-Mesery M, Amin M, et al. Diagnostic and prognostic value of hematological and immunological markers in COVID-19 infection: A meta-analysis of 6320 patients. PLoS One. 2020;15(8):e0238160.

14. Zeng F, Huang Y, Guo Y, Yin M, Chen X, Xiao L, et al. Association of inflammatory markers with the severity of COVID-19: A meta-analysis. Int J Infect Dis. 2020;96:467–74.

15. Aziz M, Fatima R, Assaly R. Elevated Interleukin-6 and Severe COVID-19: A Meta-Analysis. J Med Virol. 2020;0-1.

16. Coomes EA, Haghbayan H. Interleukin-6 in COVID-19: A Systematic Review and Meta-Analysis. *medRxiv*. 2020 Apr;2020.03.30.20048058.

17. Lagunas-Rangel FA, Chávez-Valencia V. High IL-6/IFN-γ ratio could be associated with severe disease in COVID-19 patients. J Med Virol. 2020;

18. Grifoni E, Valoriani A, Cei F, Lamanna R, Gelli AMG, Ciambotti B, et al. Interleukin-6 as prognosticator in patients with COVID-19. J Infect. 2020;81:452–82.

19. Han H, Ma Q, Li C, Liu R, Zhao L, Wang W, et al. Profiling serum cytokines in COVID-19 patients reveals IL-6 and IL-10 are disease severity predictors. Emerg Microbes Infect. 2020;9(1):1123–30.

20. Liu Z, Long W, Tu M, Chen S, Huang Y, Wang S, et al. Lymphocyte subset (CD4+, CD8+) counts reflect the severity of infection and predict the clinical outcomes in patients with COVID-19. J Infect. 2020;81:318–56.

21. Henry BM, De Oliveira MHS, Benoit S, Plebani M, Lippi G. Hematologic, biochemical and immune biomarker abnormalities associated with severe illness and mortality in coronavirus disease 2019 (COVID-19): A meta-analysis. Clin Chem Lab Med. 2020;58(7):1021–8.

22. Moher D, Liberati A, Tetzlaff J, Altman DG. Preferred reporting items for systematic reviews and meta-analyses: The PRISMA statement. Int J Surg. 2010;8(5):336–41.

23. Wan X, Wang W, Liu J, Tong T. Estimating the sample mean and standard deviation from the sample size, median, range and / or interquartile range. 2014;1-13.

24. Nie S, Zhao X, Zhao K, Zhang Z, Zhang Z, Zhang Z. Metabolic disturbances and inflammatory dysfunction predict severity of coronavirus disease 2019 (COVID-19): a retrospective study. *medRxiv*. 2020;2020.03.24.20042283.

25. Higgins JPT, Thomas J, Chandler J, Cumpston M, Li T, Page MJ WV (editors), editor. Cochrane Handbook for Systematic Reviews of Interventions version 6.0. Cochrane;

26. Zhang B, Zhou X, Zhu C, Feng F, Qiu Y, Feng J, et al. Immune phenotyping based on neutrophil-to-lymphocyte ratio and IgG predicts disease severity and outcome for patients with COVID-19. *medRxiv*. 2020;2020.03.12.20035048.

27. Viechtbauer W. Conducting meta-analyses in R with the metafor. J Stat Softw. 2010;36(3):1–48.

28. Feng X, Li P, Ma L, Liang H, Lei J, Li W, et al. Clinical Characteristics and Short-Term Outcomes of Severe Patients with COVID-19 in Wuhan, China. *medRxiv*. 2020;2020.04.24.20078063.

29. Shi H, He L, Sun W, Xu J, Wang M, Chen X, et al. Clinical Characteristics and Prognostic Factors of 148 COVID-19 Cases in a Secondary Epidemic Area. SSRN Electron J. 2020;

30. Song C-Y, Xu J, He J-Q, Lu Y-Q. COVID-19 early warning score: a multi-parameter screening tool to identify highly suspected patients. medRxiv. 2020;2020.03.05.20031906.

31. zhang huizheng, wang xiaoying, fu zongqiang, luo ming, zhang zhen, zhang ke, et al. Potential Factors for Prediction of Disease Severity of COVID-19 Patients. *medRxiv*. 2020;2020.03.20.20039818.

32. Zhao Y, Qin L, Zhang P, Li K, Liang L, Sun J, et al. Longitudinal Profiling of Cytokines and Chemokines in COVID-19 Reveals Inhibitory Mediators IL-1Ra and IL-10 Are Associated with Disease Severity While Elevated RANTES Is an Early Predictor of Mild Disease. SSRN Electron J. 2020;

33. Chen G, Wu D, Guo W, Cao Y, Huang D, Wang H. Clinical and immunologic features in severe and moderate forms of Coronavirus Disease 2019. J Clin Invest. 2020;27(1095):16–20023903.

34. He R, Lu Z, Zhang L, Fan T, Xiong R, Shen X, et al. The clinical course and its correlated immune status in COVID-19 pneumonia. J Clin Virol. 2020;127(April):104361.

35. Lv Z, Cheng S, Le J, Huang J, Feng L, Zhang B, et al. Clinical characteristics and co-infections of 354 hospitalized patients with COVID-19 in Wuhan, China: a retrospective cohort study. Microbes Infect. 2020;

36. Qin C, Zhou L, Hu Z, Zhang S, Yang S, Tao Y, et al. Dysregulation of immune response in patients with COVID-19 in Wuhan, China. Clin Infect Dis. 2020;2019(Xx Xxxx):4-10.

37. Wan S, Yi Q, Fan S, Lv J, Zhang X, Guo L, et al. Relationships among lymphocyte subsets, cytokines, and the pulmonary inflammation index in coronavirus (COVID-19) infected patients. Br J Haematol. 2020;(April):428-37.

38. Wang Z, Yang B, Li Q, Wen L, Zhang R. Clinical Features of 69 Cases with Coronavirus Disease 2019 in Wuhan, China. Clin Infect Dis. 2020;(Xx Xxxx):1-9.

39. Wei X, Su J, Yang K, Wei J, Wan H, Cao X, et al. Elevations of serum cancer biomarkers correlate with severity of COVID-19. J Med Virol. 2020;0-2.

40. Xu B, FAN C yu, WANG A lu, ZOU Y long, YU Y han, He C, et al. Suppressed T cell-mediated immunity in patients with COVID-19: A clinical retrospective study in Wuhan, China. J Infect. 2020;(xxxx).

41. Zhang J, Yu M, Tong S, Liu LY, Tang L V. Predictive factors for disease progression in hospitalized patients with coronavirus disease 2019 in Wuhan, China. J Clin Virol. 2020;127(March):104392.

42. Zheng S, Fan J, Yu F, Feng B, Lou B, Zou Q, et al. Viral load dynamics and disease severity in patients infected with SARS-CoV-2 in Zhejiang province, China, January-March 2020: Retrospective cohort study. BMJ. 2020;369(March):1-8.

43. Zhu Z, Cai T, Fan L, Lou K, Hua X, Huang Z, et al. Clinical value of immune-inflammatory parameters to assess the severity of coronavirus disease 2019. Int J Infect Dis. 2020;95:332–9.

44. National Health Commission & State Administration of Traditional Chinese Medicine. Diagnosis and Treatment Protocol for Novel Coronavirus Pneumonia. National Health Commission of the People’s Republic of China. 2020.

45. World Health Organization. (2020). Clinical management of COVID-19: interim guidance, 27 May 2020. World Health Organization. 2020.

46. Wang F, Hou H, Luo Y, Tang G, Wu S, Huang M, et al. The laboratory tests and host immunity of COVID-19 patients with different severity of illness. 2019;2(7):1–11.

47. Cifaldi L, Prencipe G, Caiello I, Bracaglia C, Locatelli F, De Benedetti F, et al. Inhibition of natural killer cell cytotoxicity by interleukin-6: Implications for the pathogenesis of macrophage activation syndrome. Arthritis Rheumatol. 2015;67(11):3037–46.

48. Liu T, Zhang J, Yang Y, Ma H, Li Z, Zhang J, et al. The role of interleukin-6 in monitoring severe case of coronavirus disease 2019. EMBO Mol Med. 2020; 12(7):e12421.

49. de Brito R de CCM, Lucena-Silva N, Torres LC, Luna CF, Correia J de B, da Silva GAP. The balance between the serum levels of IL-6 and IL-10 cytokines discriminates mild and severe acute pneumonia. BMC Pulm Med. 2016;16(1):19–21.

50. Chen Y, Rubin P, Williams J, Hernady E, Smudzin T, Okunieff P. Circulating IL-6 as a predictor of radiation pneumonitis. Int J Radiat Oncol Biol Phys. 2001;49(3):641–8.

51. Diao B, Wang C, Tan Y, Chen X, Liu Y, Ning L, et al. Reduction and Functional Exhaustion of T Cells in Patients With Coronavirus Disease 2019 (COVID-19). Front Immunol. 2020;11(May):1-7.

52. Moore KW, Malefyt RDW, Robert L, Garra AO. Interleukin-10 and the Interleukin-10 Receptor. Mol Cell Biol. 2001;1(1):683–765.

53. Maris CH, Chappell CP, Jacob J. Interleukin-10 plays an early role in generating virus-specific T cell anergy. BMC Immunol. 2007;8:9–12.

54. Brooks DG, Trifilo MJ, Edelmann KH, Teyton L, McGavern DB, Oldstone MBA. Interleukin-10 determines viral clearance or persistence in vivo. Nat Med. 2006;12(11):1301–9.

55. Ejrnaes M, Filippi CM, Martinic MM, Ling EM, Togher LM, Crotty S, et al. Resolution of a chronic viral infection after interleukin-10 receptor blockade. J Exp Med. 2006;203(11):2461–72.

56. Biswas PS, Pedicord V, Ploss A, Menet E, Leiner I, Pamer EG. Pathogen-Specific CD8 T Cell Responses Are Directly Inhibited by IL-10. J Immunol. 2007;179(7):4520–8.

57. Yang Z, Liu J, Zhou Y, Zhao X, Zhao Q, Liu J. The effect of corticosteroid treatment on patients with coronavirus infection: a systematic review and meta-analysis. J Infect. 2020;81:13–20.

58. Stallmach A, Kortgen A, Gonnert F, Coldewey SM, Reuken P, Bauer M. Infliximab against severe COVID-19-induced cytokine storm syndrome with organ failure – A cautionary case series. Vol. 24, Critical Care. BioMed Central; 2020. p. 444.

59. Duret PM, Sebbag E, Mallick A, Gravier S, Spielmann L, Messer L. Recovery from COVID-19 in a patient with spondyloarthritis treated with TNF-αlpha inhibitor etanercept. Annals of the Rheumatic Diseases. BMJ Publishing Group; 2020.

60. Tursi A, Angarano G, Monno L, Saracino A, Signorile F, Ricciardi A, et al. COVID-19 infection in Crohn’s disease under treatment with adalimumab. Vol. 69, Gut. BMJ Publishing Group; 2020. p. 1364-5.

61. Bezzio C, Manes G, Bini F, Pellegrini L, Saibeni S. Infliximab for severe ulcerative colitis and subsequent SARS-CoV-2 pneumonia: a stone for two birds. Gut. NLM (Medline); 2020.

62. Jamilloux Y, Henry T, Belot A, Viel S, Fauter M, El Jammal T, et al. Should we stimulate or suppress immune responses in COVID-19? Cytokine and anti-cytokine interventions. Autoimmun Rev. 2020;(April):102567.

63. Kalechman Y, Gafter U, Gal R, Rushkin G, Yan D, Albeck M, et al. Anti-IL-10 Therapeutic Strategy Using the Immunomodulator AS101 in Protecting Mice from Sepsis-Induced Death: Dependence on Timing of Immunomodulating Intervention. J Immunol. 2002;169(1):384–92.

64. Nalbant A, Kaya T, Varim C, Yaylaci S, Tamer A, Cinemre H. Can the neutrophil/lymphocyte ratio (NLR) have a role in the diagnosis of coronavirus 2019 disease (COVID-19)? Rev Assoc Med Bras. 2020;66(6):746–51.

65. Yufei Y, Mingli L, Xuejiao L, Xuemei D, Yiming J, Qin Q, et al. Utility of the neutrophil-to-lymphocyte ratio and C-reactive protein level for coronavirus disease 2019 (COVID-19). Scand J Clin Lab Invest. 2020;0(0):1–5.

66. Liu Y, Du X, Chen J, Jin Y, Peng L, Wang HHX, et al. Neutrophil-to-lymphocyte ratio as an independent risk factor for mortality in hospitalized patients with COVID-19. J Infect. 2020 Jul;81(1):e6-12.

67. Huang I, Pranata R, Lim MA, Oehadian A, Alisjahbana B. C-reactive protein, procalcitonin, D-dimer, and ferritin in severe coronavirus disease-2019: a meta-analysis. Ther Adv Respir Dis. 2020;14:1–14.

68. Soraya GV, Ulhaq ZS. Crucial laboratory parameters in COVID-19 diagnosis and prognosis: An updated meta-analysis. Med Clin (Barc). 2020 Aug;155(4):143–51.

69. Shen B, Yi X, Sun Y, Liu H, Chen H, Guo T, et al. Article Proteomic and Metabolomic Characterization of COVID-19 Patient Sera ll Article Proteomic and Metabolomic Characterization of COVID-19 Patient Sera. 2020;59-72.

